# Recurrent and Concurrent Prediction of Longitudinal Progression of Stargardt Atrophy and Geographic Atrophy

**DOI:** 10.1101/2024.02.11.24302670

**Authors:** Zubin Mishra, Ziyuan Wang, Emily Xu, Sophia Xu, Iyad Majid, SriniVas R. Sadda, Zhihong Jewel Hu

**Author notes:** Correspondence and requests: Zhihong Jewel Hu, Doheny Eye Institute, 150 North Orange Grove Blvd, Pasadena, CA 91103.

## Abstract

Stargardt disease and age-related macular degeneration are the leading causes of blindness in the juvenile and geriatric populations, respectively. The formation of atrophic regions of the macula is a hallmark of the end-stages of both diseases. The progression of these diseases is tracked using various imaging modalities, two of the most common being fundus autofluorescence (FAF) imaging and spectral-domain optical coherence tomography (SD-OCT). This study seeks to investigate the use of longitudinal FAF and SD-OCT imaging (month 0, month 6, month 12, and month 18) data for the predictive modelling of future atrophy in Stargardt and geographic atrophy. To achieve such an objective, we develop a set of novel deep convolutional neural networks enhanced with recurrent network units for longitudinal prediction and concurrent learning of ensemble network units (termed ReConNet) which take advantage of improved retinal layer features beyond the mean intensity features. Using FAF images, the neural network presented in this paper achieved mean (± standard deviation, SD) and median Dice coefficients of 0.895 (± 0.086) and 0.922 for Stargardt atrophy, and 0.864 (± 0.113) and 0.893 for geographic atrophy. Using SD-OCT images for Stargardt atrophy, the neural network achieved mean and median Dice coefficients of 0.882 (± 0.101) and 0.906, respectively. When predicting only the interval growth of the atrophic lesions with FAF images, mean (± SD) and median Dice coefficients of 0.557 (± 0.094) and 0.559 were achieved for Stargardt atrophy, and 0.612 (± 0.089) and 0.601 for geographic atrophy. The prediction performance in OCT images is comparably good to that using FAF which opens a new, more efficient, and practical door in the assessment of atrophy progression for clinical trials and retina clinics, beyond widely used FAF. These results are highly encouraging for a high-performance interval growth prediction when more frequent or longer-term longitudinal data are available in our clinics. This is a pressing task for our next step in ongoing research.

## 1 Introduction

Stargardt disease is a recessive inherited disorder that is the most common form of juvenile-onset macular dystrophy, causing progressive damage or degeneration of the macula [1-14]. AMD is the leading cause of blindness in people aged 65 and older in the western world, with many of these patients appearing to eventually lose vision due to the development of macular neovascularization and non-neovascularization or geographic atrophy (GA). Stargardt atrophy and geographic atrophy are the end points of Stargardt disease and non-neovascularization AMD respectively.

The atrophic formation in juvenile and age-related macular dystrophy is the most severe cause of vision loss and blindness. Until recently, there had been no proven effective treatments for macular atrophy. Currently, two complement inhibitors, pegcetocoplan (Apellis Pharmaceuticals) and avacincaptad (Iveric Bio), demonstrated positive Phase 3 clinical trial results in treating AMD atrophy and were cleared for clinic use by the Food and Drug Administration (FDA). As such, detecting macular atrophy and predicting the expected progression, and selecting the optimal therapy have been topics of pressing need and critical importance.

Fundus autofluorescence (FAF) imaging and spectral-domain optical coherence tomography (SD-OCT) are two widely accessible imaging modalities that can aid in the diagnosis and monitoring of both diseases [15]. FAF imaging provides an in vivo assay of the lipofuscin content within the retinal pigment epithelium (RPE) cells, while SD-OCT allows for the three-dimensional visualization of the retina’s microstructure and the direct evaluation of individual retinal layers, including the photoreceptors and RPE [16-17].

Approaches to the automated analysis of Stargardt disease and geographic atrophy frequently make use of convolutional neural network (CNN) architecture for semantic segmentation [18-37]. However, current approaches to automated analysis for macular atrophy generally do not incorporate longitudinal data, focusing on one instant in time. One way to incorporate longitudinal data is through the use of recurrent neural networks, such as the long short-term memory (LSTM) architecture, a modification of the recurrent neural network [38-45].

The LSTM architecture has been used in medical image analysis for the detection of cardiomegaly, consolidation, pleural effusion and hiatus hernia on X-ray and for Alzheimer’s disease diagnosis [46-47]. In the field of ophthalmology, LSTMs have been successfully used for assessing glaucoma progression, predicting diabetic retinopathy, and predicting late AMD progression. However, in these cases, the LSTM architecture was used for classification, not segmentation, of images. Moreover, the retinal layers in volumetric SD-OCT images include rich information beyond the generally used image features (original retinal layer mean intensity and thickness features). To take advantage of the rich 3D retinal layer information, an extension of CNNs to address this issue could be ensemble CNNs, where multiple neural networks are used together to each handle different input of the novel features based on the retinal layer and concurrently predict the output of atrophy progression with the CNN features from the ensemble networks together [18, 29-30]. This study seeks to investigate the predictive modelling of future progression of atrophy in Stargardt and geographic atrophy based on longitudinal data from multiple patients’ visits using novel deep convolutional neural networks (termed ReConNet) enhanced with convolutional recurrent network (i.e., LSTM in this project) units and concurrent learning of ensemble network units. Our major contributions include: 1) The novel ReConNet architecture incorporates LSTM connections with encoder and decoder pathways for use on longitudinal data. 2) It takes advantage of the enhanced OCT en face feature maps in an ensemble network to capture higher order inherent retinal layer features associated with atrophy for enhanced algorithm performance. 3) It can be used for the generation or prediction for both future FAF images and future Stargardt atrophy and geographic atrophy.

## 2 Materials and Methods

### 2.1 Imaging Dataset and Ground Truth

141 eyes from 100 patients diagnosed with Stargardt disease were identified from the ProgStar study that had FAF imaging (Spectralis HRA+OCT; Heidelberg Engineering) performed at initial (zero-month) visit and six-, twelve- and eighteen-month follow-ups [1-4]. Of these eyes 71 were identified that additionally had SD-OCT imaging (Spectralis HRA+OCT; Heidelberg Engineering) performed at initial baseline and six-month visits. The geographic atrophy dataset consisted of 60 eyes from 60 patients obtained from Doheny Eye Institute clinics that had FAF imaging performed at initial baseline visit and six-, twelve- and eighteen-month follow-ups. All of the FAF images were manually segmented by certified graders to create ground truth masks marking the atrophied regions of the retina. 85% of the images were used for training, and 15% for testing. For the geographic atrophy dataset, the field of view was 30° with pixel dimensions of 768 × 868. For the Stargardt disease dataset, the field of view was 20° with pixel dimensions of 512 × 512. The FAF image scan dimensions are resized to a standard size of 256 × 256. The SD-OCT image scan dimensions are 496 (depth) × 1024 (A-scans) × 49 (B-scans) pixels or 496 (depth) × 512 (A-scans) × 49 (B-scans) pixels. The 496 × 512 images were resized to a standard width of 1024 and standard height of 496. After segmentation, 49 × 1024 feature maps were produced from each SD-OCT volume. These feature maps were resized to 512 × 512 for registration to FAF images and later resized to 256 × 256 prior to being input to the neural network due to memory constraints. All the right (OD) eye SD-OCT scans and FAF images were flipped horizontally to provide consistency in the analysis. As described in our previous work, all Stargardt dataset FAF images and SD-OCT-derived feature maps were registered to the initial baseline visit through feature-based image registration [48]. All geographic atrophy dataset FAF images were similarly registered to their initial visit images. For Stargardt FAF images, contrast limited adaptive histogram equalization was performed as part of pre-processing.

The ground truths utilized in this study for both geographic atrophy and Stargardt atrophy were based on FAF images, as thus far FAF images have been utilized as standard imaging modality for macular atrophy assessment in clinical trial studies. Geographic atrophy and Stargardt atrophy lesions on FAF images were graded using the semiautomated software tool RegionFinder (Heidelberg Engineering). Images were initially graded by a certified reading center grader, and the grading is subsequently reviewed by a senior grader. A senior investigator (SS) resolved discrepancies between the two graders.

### 2.2 Neural Network Structure

In this project, we designed three network structures (ReConNet, ReConNet-Ensemble, ReConNet-Interval) for three practical atrophy prediction applications based on multiple patient visits: 1) Prediction of future geographic atrophy and Stargardt atrophy regions using longitudinal FAF images (ReConNet); 2) Prediction of future Stargardt atrophy regions using longitudinal SD-OCT images (ReConNet-Ensemble); 3) Prediction of interval growth of geographic atrophy and Stargardt atrophy regions using longitudinal FAF images (ReConNet-Interval). The algorithms were implemented using the open-source deep learning framework Keras.

The first neural network, ReConNet, used in this study is for longitudinal prediction of progression of geographic atrophy and Stargardt atrophy on FAF images based on the network architecture which has an encoding pathway and a decoding pathway, with a concatenation linking the output of an encoding pathway convolution to the input of a decoding pathway convolution. The term ReConNet is used for both FAF and OCT to be flexible in the future to include additional ensemble network inputs for concurrent learning. Such a deep learning architecture can be used for semantic segmentation using a relatively small training set of images [18]. To incorporate longitudinal data, the neural network in this study has multiple encoding pathways, one for each timestep. At each layer of the encoding pathway, the outputs of the encoding pathway convolutions are combined using a 2D convolutional-LSTM layer. The output of this layer is concatenated with the input of the corresponding decoding pathway convolution. Note that the overall network architectures for ReConNet and ReConNet-Interval are similar, and the difference is that the input for ReConNet is original FAF image (and related ground truth) and for ReConNet-Interval is the FAF image with the region of atrophy set to zero (and related interval ground truth) reflecting the growth regions between any two adjacent patient visits. A schematic of the neural network used in this study is depicted in Figure 1.

**Figure 1.**
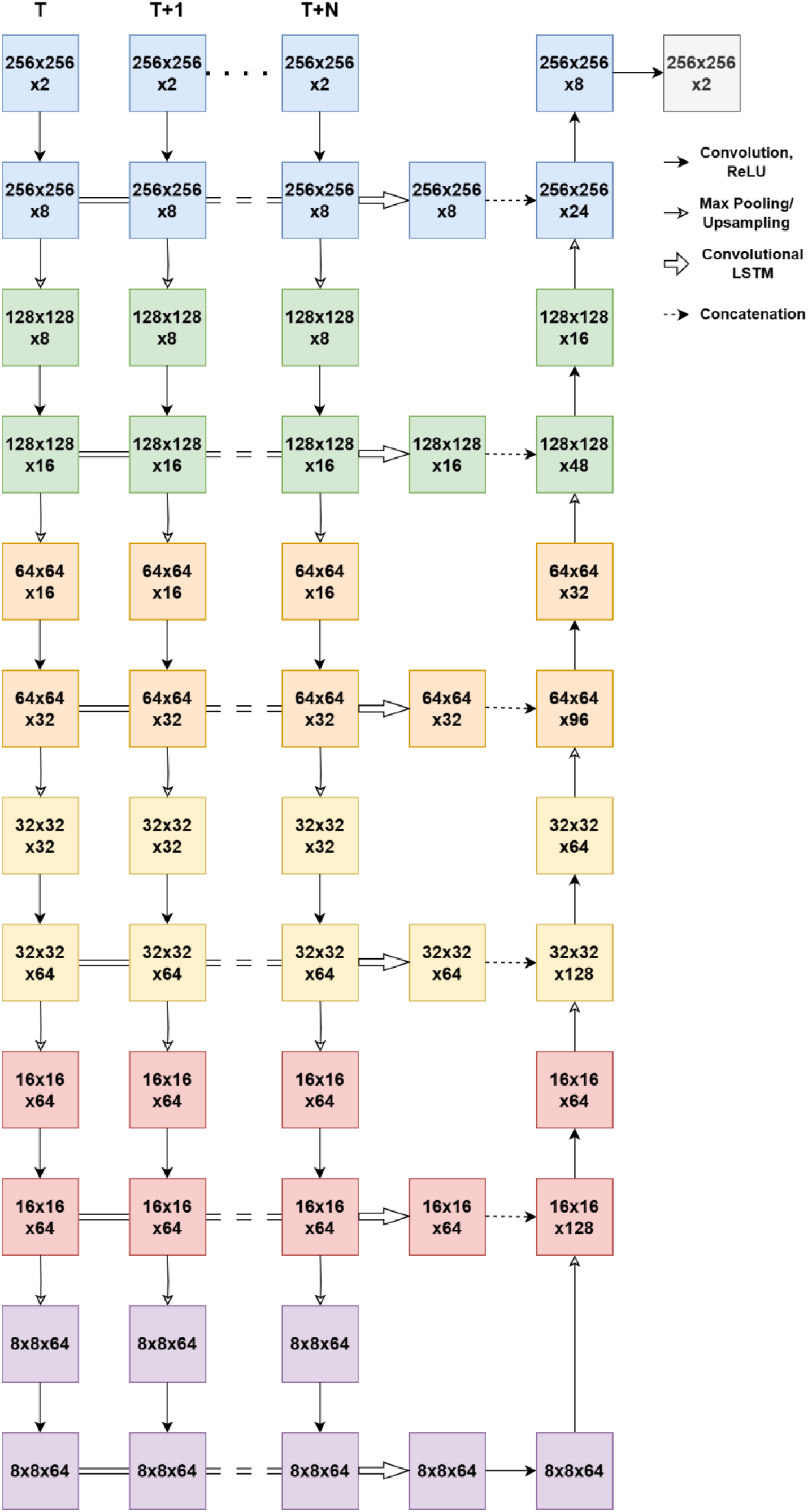
Schematic of the ReConNet neural network architecture.

The second neural network structure, ReConNet-Ensemble, used in this study is for longitudinal prediction of progression of Stargardt atrophy on OCT images. The ensemble inputs and network structure of ReConNet-Ensemble are similar to that previously described by our team [18]. The difference is that the previous one is for prediction using baseline alone and the ReConNet-Ensemble in this project includes longitudinal data with three patient visits (month 0, month 6, and month 12) reflecting dynamic changes of atrophy over time. In ReConNet-Ensemble neural network, multiple of the above-described neural network are used to take in several OCT-derived feature maps as input, allowing for the incorporation of the three-dimensional OCT data. These feature maps are en face images derived from the OCT data surrounding the ellipsoid zone in the depth (z-direction) of the OCT, the region of the retina most affected by Stargardt atrophy. These feature maps also extend beyond the traditional mean intensity maps, additionally including minimum intensity, maximum intensity, median intensity, standard deviation, skewness, kurtosis, gray level entropy, and thickness of the ellipsoid zone. In our previous work, we found significant improvement for the prediction of Stargardt atrophy when incorporating these advanced feature maps compared to using mean intensity alone. The logits layers of the individual neural networks are combined through averaging, with the combined result subsequently being sent through a softmax function to predict probability maps of atrophy. The ReConNet-ensemble neural network is depicted in Figure 2.

**Figure 2.**
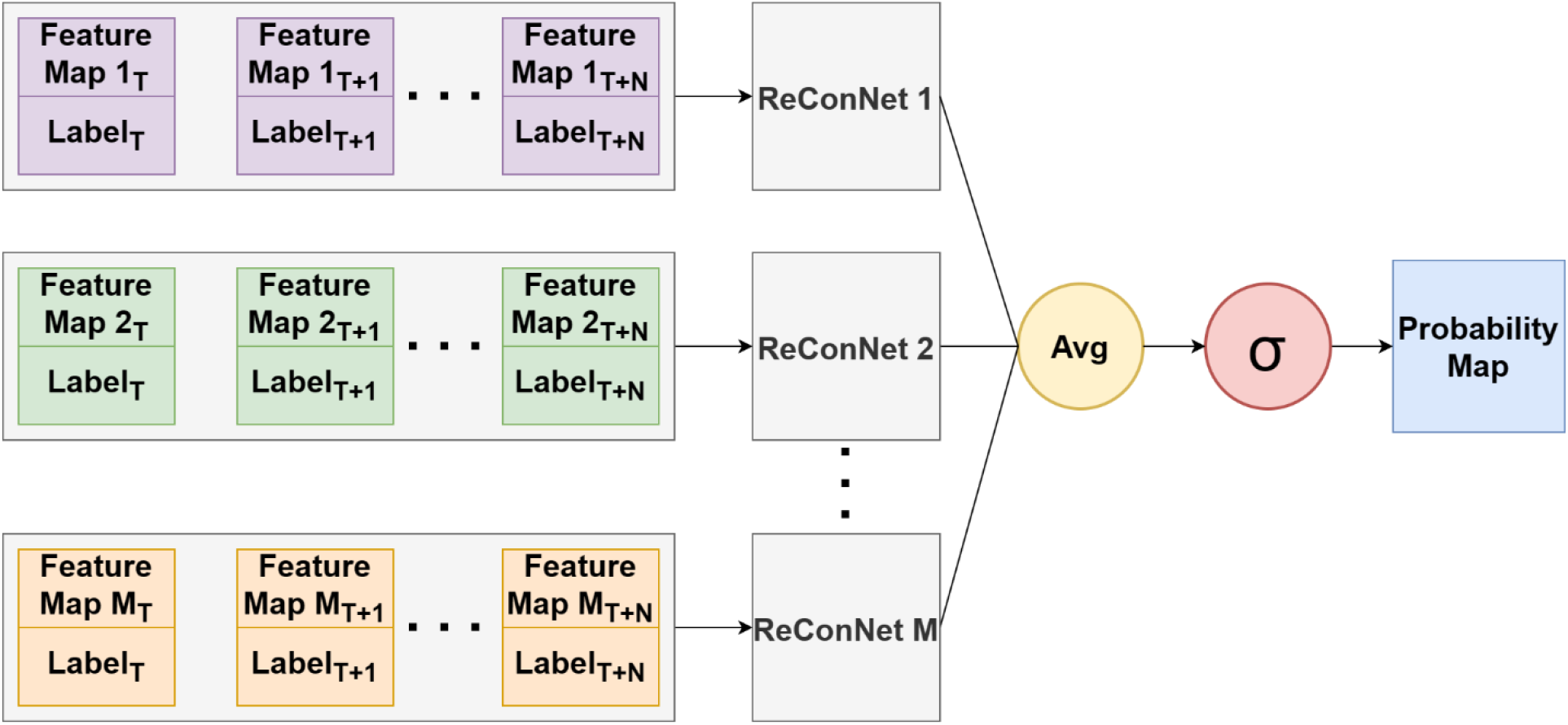
Ensemble neural network structure. The OCT feature maps include traditional mean intensity maps, and additional minimum intensity, maximum intensity, median intensity, standard deviation, skewness, kurtosis, gray level entropy, and thickness of the ellipsoid zone.

Note that the ReConNet neural networks can predict both future images and atrophy. They are optimized by different loss functions depending on if used for image generation or for segmentation. For image generation, cosine similarity loss is used:

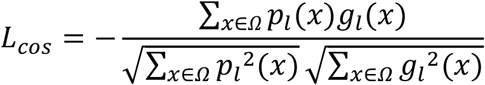

For segmentation, Dice loss is used:

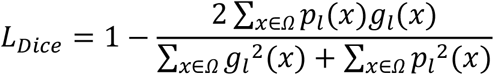

In the above equations *g*_*l*_ (*x*) is the ground truth probability the pixel *x* ∈ *Ω* belongs to class *l, p*_*l*_ (*x*) is the estimated probability that pixel *x* belongs to class *l*.

### 2.3 Atrophy Prediction

#### 2.3.1 Prediction of future geographic atrophy and Stargardt atrophy regions using longitudinal FAF images (ReConNet)

The prediction of progression of geographic and Stargardt atrophy using longitudinal FAF images is trained with two steps (initial step of ReConNet1 and final step of ReConNet2. See Figure 3 for details). The initial step consists of the prediction/generation of FAF image and atrophy regions (size and location) on the FAF image. The input for the neural network was the zero-month, six-month, and twelve-month FAF images, each paired with the manually graded atrophy label for that visit. The outputs were the predicted/generated FAF image and atrophy at eighteen-months. In the second step, the generated/predicted FAF image and label are then used in conjunction with the zero-month, six-month, and twelve-month images in the neural network for enhanced atrophy prediction. An example of this process with the input and output at each step is shown in Figure 3.

**Figure 3.**
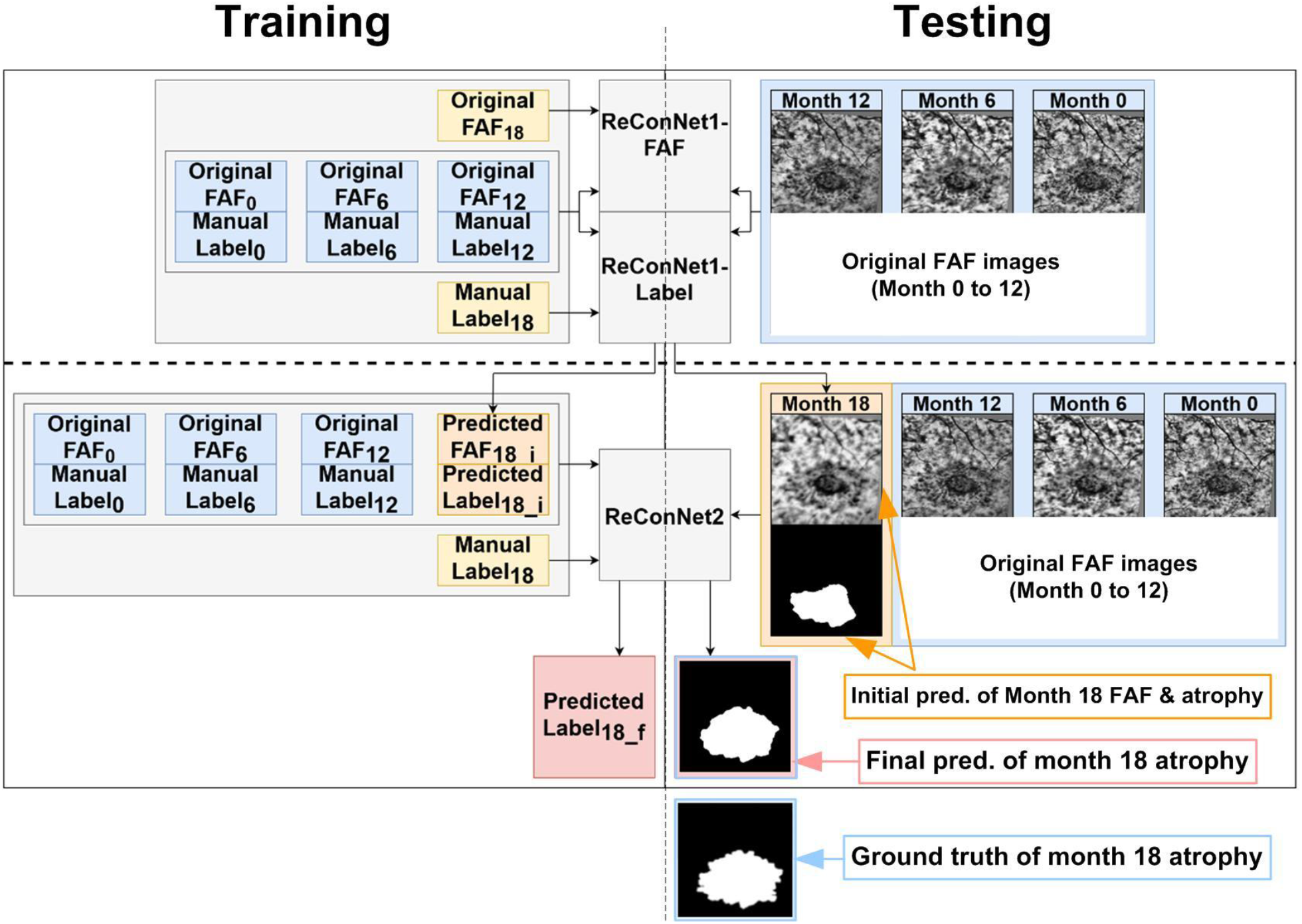
Schematic of the prediction algorithm, where i in the subscript indicates the initial prediction results from ReconNet1 and f in the subscript indicates the final prediction results from ReConNet2.

#### 2.3.2 Prediction of progression of Stargardt atrophy using longitudinal SD-OCT images (ReConNet-Ensemble)

With the ensemble neural network for Stargardt atrophy progression using longitudinal SD-OCT images, a similar process of two steps (ReConNet1-Ensemble and ReConNet2-Ensemble) is done. Note that for OCT data, we only have data from three patient visits (month 0, month 6, month 12) Instead of using single input of FAF image for each patient visit, in step 1, various enhanced feature maps at each patient visit derived from the zero-month and six-month SD-OCT scans (each again paired with the manually graded atrophy label for that visit), are sent through the ensemble network, producing both a predicted region of atrophy and a predicted en face image. As mentioned, the enhanced features demonstrate rich 3D atrophy information beyond the mean intensity associated features as shown in our previous paper [18]. In step 2, rather than attempt to predict the individual twelve-month OCT-derived feature maps, the predicted en face image is used as a stand-in for the twelve-month feature maps and sent through the neural network again in conjunction with the zero-month and six-month feature maps.

#### 2.3.3 Prediction of interval growth of geographic atrophy and Stargardt atrophy regions using longitudinal FAF images (ReConNet-Interval)

In this task, the interval growth of geographic atrophy and Stargardt atrophy between visits was examined. In the zero-month, six-month, and twelve-month FAF images, the areas labeled as atrophy by graders was set to zero, and interval growth labels were paired with each visit. A placeholder label created by inverting the twelve-month atrophy region ground truth was paired with the twelve-month FAF image. The neural network, ReConNet-Interval was trained to identify only the interval growth of the region of atrophy. Note that different from ReConNet and ReConNet-Ensemble which are performed by two steps where the generated/predicted image and label are used in conjunction with the zero-month, six-month, and twelve-month images in the neural network for enhanced atrophy prediction, the ReConNet-Interval can only be performed for the first step, as the second step will need an additional visit after 18-month to obtain the interval. Hence the performance of the one step ReConNet1-Interval may not be comparable with the enhanced atrophy prediction results after two steps from ReConNet2 and ReConNet2-Ensemble.

### 2.4 Performance Evaluation

Four metrics are used to evaluate the performance of the neural networks: pixel-wise accuracy, Dice coefficient, sensitivity, and specificity. Pixel-wise accuracy measures all correctly identified pixels in the image:

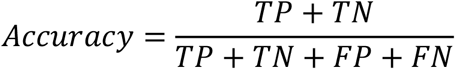

Where TP is true positives, TN is true negatives, FP is false positives, and FN is false negative. Sensitivity describes how much atrophy is correctly labeled compared to the total amount of atrophy:

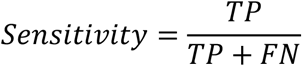

Specificity describes how much non-atrophied tissue is correctly labeled compared to the total amount of non-atrophied tissues:

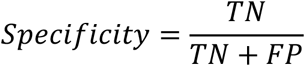

The Dice coefficient of two sets A and B describes the spatial overlap between the two sets:

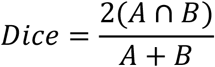

## 3 Results

Neural network predictions were evaluated using the pixel-wise accuracy, Dice coefficient, sensitivity, and specificity. The initial and final prediction results for ReConNet are shown in Table 1. The initial and final results for ReConNet-Ensemble are shown in Table 2. The results for ReConNet-Interval are shown in Table 3.

**Table 1.** Results for ReConNet (Median|Mean(SD))

|  |  | Accuracy | Dice Coefficient | Sensitivity | Specificity |
| --- | --- | --- | --- | --- | --- |
| Stargardt Atrophy | ReConNet1-Initial | 0.919\|0.904(0.072) | 0.568\|0.577(0.163) | 0.406\|0.432(0.163) | 0.998\|0.996(0.008) |
|  | ReConNet2-Final | 0.98\|0.973(0.021) | 0.922\|0.895(0.086) | 0.876\|0.84(0.12) | 0.998\|0.996(0.007) |
|  | *p*-value | <0.001 | <0.001 | <0.001 | 0.020 |
| Geographic Atrophy | ReConNet1-Initial | 0.901\|0.896(0.06) | 0.867\|0.827(0.129) | 0.84\|0.839(0.107) | 0.971\|0.938(0.089) |
|  | ReConNet2-Final | 0.928\|0.919(0.042) | 0.893\|0.864(0.113) | 0.964\|0.945(0.062) | 0.92\|0.901(0.077) |
|  | *p*-value | <0.001 | <0.001 | <0.001 | <0.001 |

**Table 2.** Results for ReConNet-Ensemble (Median/Mean (SD))

|  |  | Accuracy | Dice Coefficient | Sensitivity | Specificity |
| --- | --- | --- | --- | --- | --- |
| Stargardt Atrophy | ReConNet1-Ensemble-Initial | 0.955\|0.92(0.093) | 0.742\|0.662(0.238) | 0.691\|0.63(0.292) | 0.992\|0.977(0.032) |
|  | ReConNet2-Ensemble-Final | 0.98\|0.968(0.033) | 0.906\|0.882(0.101) | 0.912\|0.894(0.086) | 0.991\|0.983(0.029) |
|  | *p*-value | <0.001 | <0.001 | <0.001 | 0.140 |

**Table 3.**
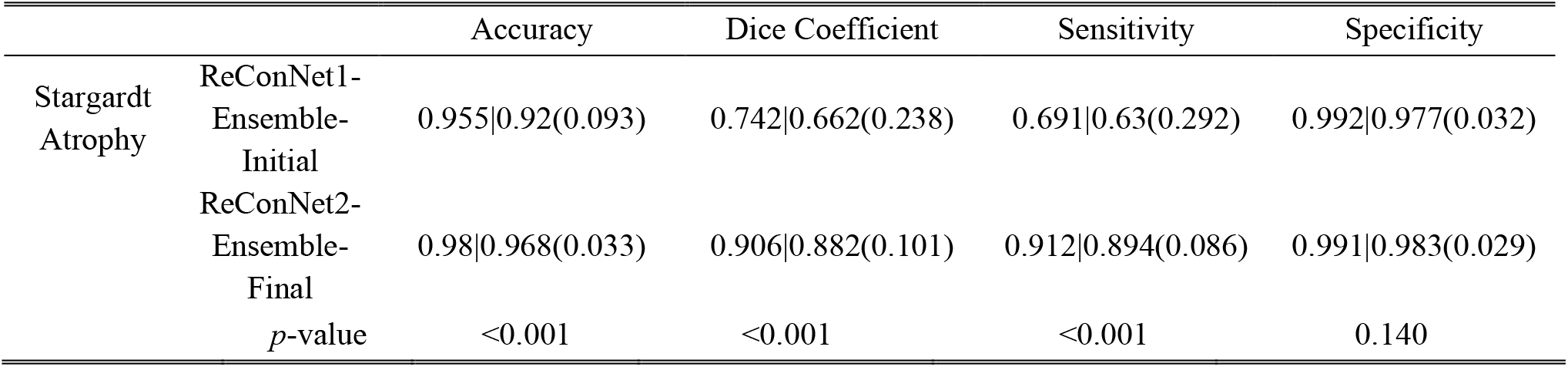
Results for ReConNet-Interval (Median/Mean (SD))

When comparing the performance of the neural network with and without the inclusion of a predicted eighteen-month FAF image, the Wilcoxon ranked-sign test was used due to the non-normal distribution of the data, resulting in p-values less than 0.05 for all metrics for all neural network configurations for both Stargardt atrophy and GA, except for with specificity in one case. Examples of inputs, output predicted regions of atrophy, and ground truths after ReConNet, ReConNet-Ensemble, and ReConNet-Interval are shown in Figure 4, 5, and 6, respectively.

**Figure 4.**
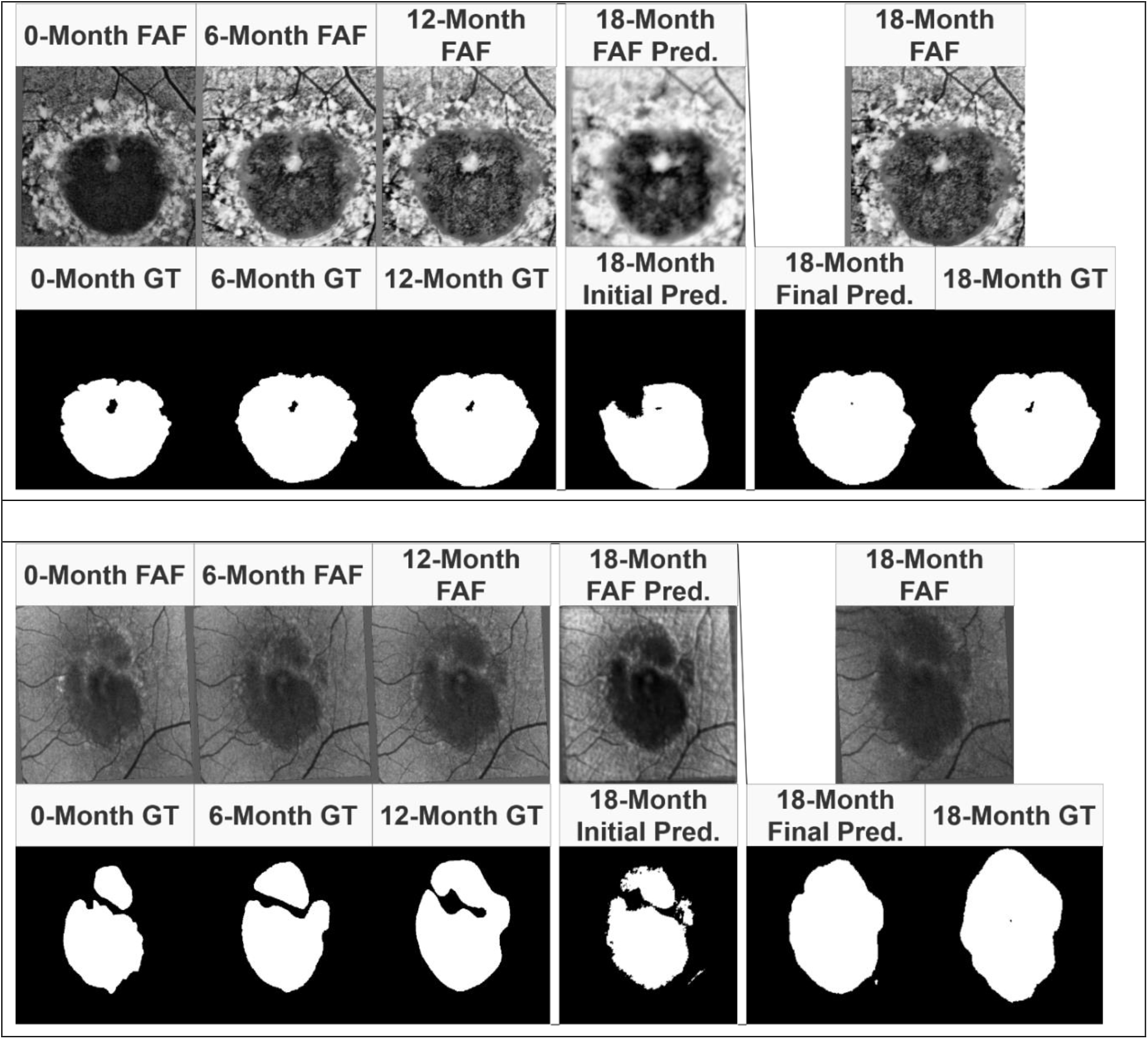
Example results of ReConNet. Input FAF images and labels, initial prediction, final prediction, and ground truth comparison for ReConNet with Stargardt atrophy (Top) and geographic atrophy (Bottom).

**Figure 5.**
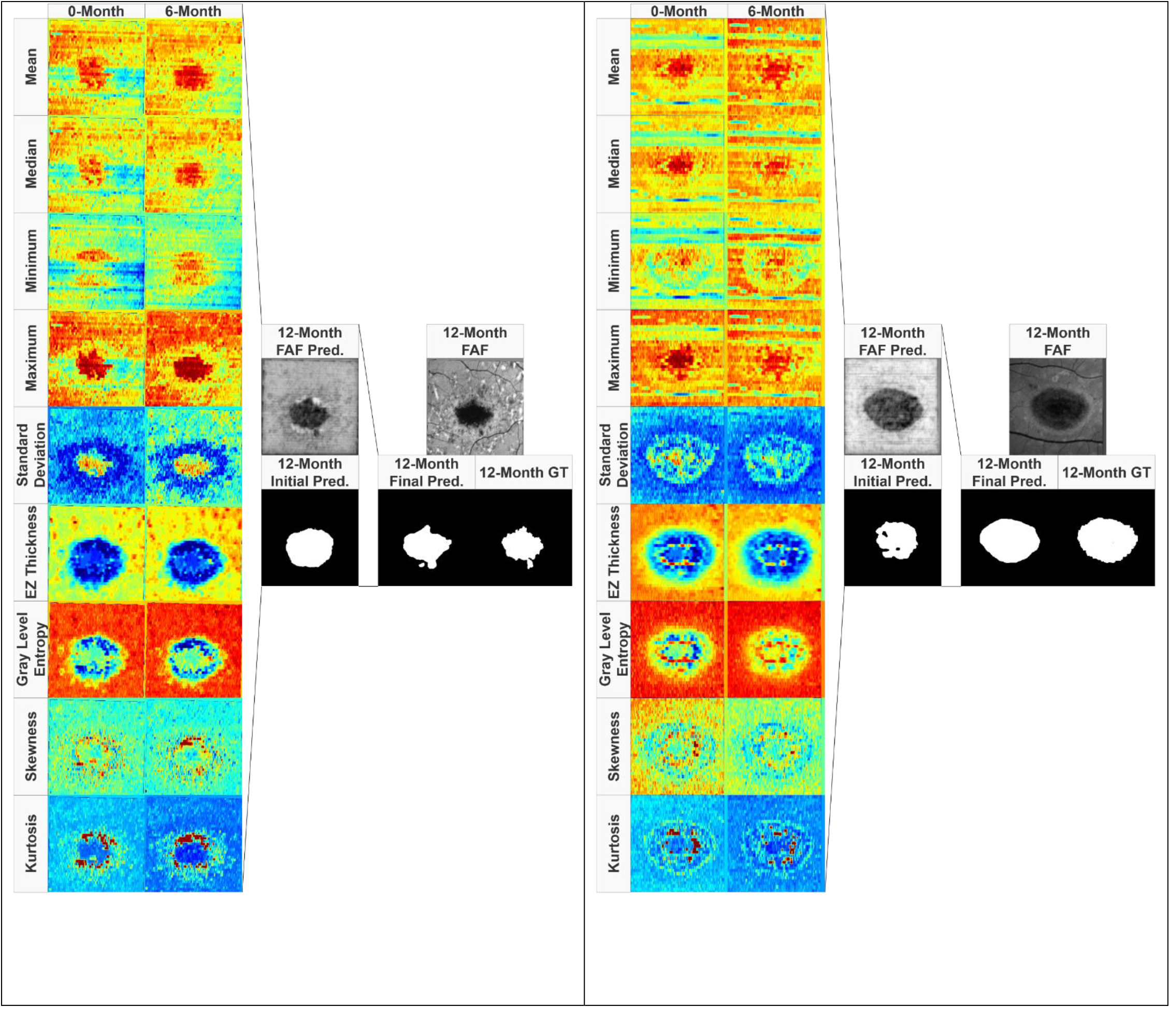
Example results after ReConNet-Ensemble. Input OCT feature maps, initial prediction, final prediction, and ground truth comparison for ensemble ReConNet with Stargardt atrophy. Input labels are not pictured.

**Figure 6.**
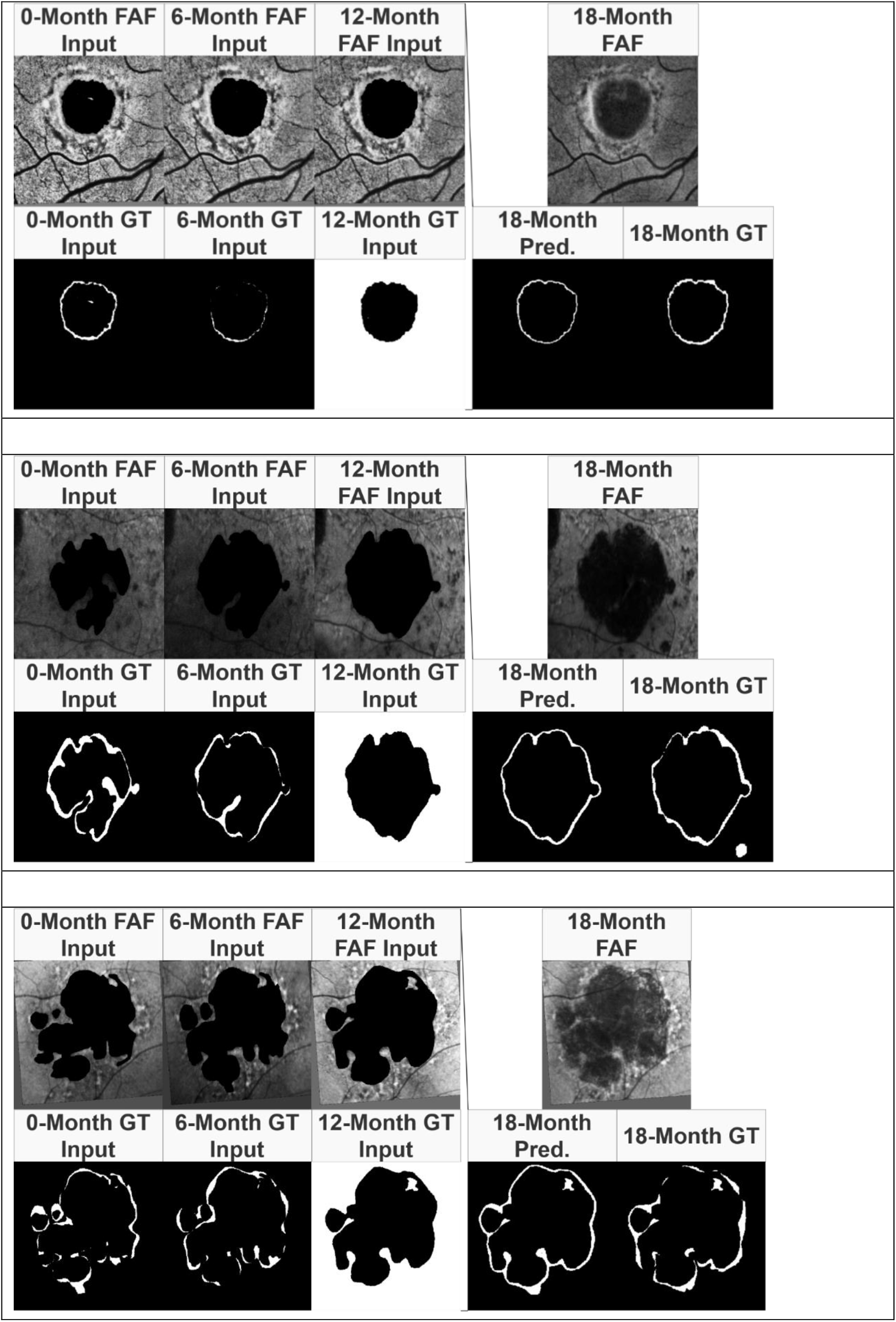
Example results after ReConNet1-Interval. (Top) Input modified FAF images, labels, interval growth prediction, and ground truth comparison with Stargardt atrophy. (Middle and Bottom) Input modified FAF images, labels, interval growth prediction, and ground truth comparison with geographic atrophy. 18-Month FAF images are shown for reference.

## 4 Discussion and Conclusions

This paper represents the first attempt to longitudinal imaging data to predict future regions of atrophy in the setting of both AMD and Stargardt disease using a recurrent and concurrent neural network architecture.

Particularly for Stargardt atrophy progression prediction, the only one other attempt to predict the progression of Stargardt disease using deep learning methods also comes from our group, in which a self-attended U-Net was developed to predict the progression of atrophy twelve months from baseline FAF images alone.

For the prediction of atrophy growth in FAF using ReConNet, as shown in Table 1, the initial ReConNet1 neural network presented in this paper achieved a median Dice coefficient of 0.568 for eighteen-month predictions of atrophy in the setting of Stargardt atrophy using baseline zero-month, six-month, and twelve-month data. When this initial generated/predicted eighteen-month images and labels was used in conjunction with prior visits of longitudinal data, a median Dice coefficient of 0.922 for ReConNet2 was achieved, showing significant improvement (62% improvement, p<0.05). For geographic atrophy, median Dice coefficients of 0.867 for ReConNet1 and 0.893 for ReConNet2 were achieved with modest enhancement, probably due to the good performance of ReConNet1 and additionally the smaller size of the geographic atrophy dataset (60 eyes) compared with the Stargardt atrophy dataset (141 eyes), hence the improvement space is limited.

For the prediction of Stargardt atrophy growth in OCT, the ensemble neural network with various OCT feature maps resulted in a similar pattern of performance. Note that the ensemble neural network was only applied on Stargardt atrophy prediction (with month 0 and 6 to predict month 12 since further longitudinal data were not available). Additionally, the longitudinal OCT data for geographic atrophy patients were not accessible in this study. Using initial zero-month and six-month data, the ensemble neural network achieved a median Dice coefficient of 0.742 for twelve-month predictions in the setting of Stargardt disease (71 eyes) for initial ReConNet1-Ensemble, and using the initial prediction in conjunction with the prior visits of longitudinal data resulted in a median Dice coefficient of 0.906 for ReConNet2-Ensemble, again showing significant improvement (22%improvement, p<0.05). Similar patterns of results were seen with pixel accuracy, sensitivity, and specificity, with the only exception of specificity for the prediction of Stargardt atrophy when using FAF images. In comparison to our previous work, we have achieved higher Dice coefficients for atrophy prediction through the inclusion of longitudinal data.

It is worth noting that the prediction results of Dice coefficient for Stargardt atrophy using OCT based on ReConNet-Ensemble (Table 2 and Figure 5) is comparable to that using FAF based on ReConNet (Table 1 and Figure 4). This finding is critically important. Usually, FAF has been considered as the best imaging modality regarding imaging quality and has been used as a main imaging modality for clinical trials of new atrophy drugs. However, the FAF imaging acquisition process is very uncomfortable for patients, and it is in two-dimensional imaging modality. OCT imaging is much tolerable for patients, is a three-dimensional imaging modality, and is becoming the most popular imaging modality in major retina clinics. Hence, our novel recurrent and concurrent neural networks, based on multiple advanced en face OCT maps which reflect higher order inherent retinal layer features associated with atrophy, provides a new and more efficient, and practical way for the prediction of atrophy progression in clinical trials and clinics.

When predicting only the interval growth of the atrophic lesions in FAF using ReConNet-Interval, in the setting of Stargardt atrophy, a median Dice coefficient of 0.557 was achieved. For geographic atrophy, a median Dice coefficient of 0.601 was achieved. As mentioned, the ReConNet-Interval can only be performed for the first step, as the second step will need an additional visit after 18 months to obtain the interval which is not available for this project. Based on our investigation of the two steps’ schema for ReConNet and ReConNet-Ensemble, we can reasonably expect that the performance of median Dice coefficient for the interval growth of Stargardt and geographic atrophic lesions for ReConNet2-Interval should be both greater than 0.80.

In Figure 4, it is possible to compare the initial and final predictions of the ReConNet neural network to the input FAF images and ground truths. Of interest in these images is improved detail of the final prediction compared to the initial prediction. This indicates that in predicting the FAF image, the neural network may capture features important to growth of atrophy that are not captured by segmentation alone. In Figure 5, it is possible to compare the initial and final predictions of the ensemble neural network based on OCT to the input feature maps and ground truths. Once again, there is an improved final prediction after the incorporation of an initially predicted FAF image and ground truth. This indicates that the benefit of incorporating a predicted FAF image for the final prediction is consistently present across imaging modalities.

In Figure 6, it is possible to compare the predicted interval growth to the input edited FAF images. It can be seen that the predicted interval growth tends to over-estimate the ground truth. This is also apparent in the high sensitivity and relatively low specificity.

This study is not without limitations. Firstly, while our ReConNet architecture performs well with limited sample size, the datasets sizes for both Stargardt disease and geographic atrophy are relatively small, particularly for the geographic atrophy dataset Secondly, the longitudinal patient visits are limited, especially for the goal for the evaluation of growth-interval. Either more longitudinal patient visits or denser visits for ReConNet-Interval should be included to achieve a reasonably good atrophy progression prediction by applying two steps of ReConNet-Interval (i.e., ReConNet1-Interval and ReConNet2-Interval) similar as the ReConNet and ReConNet-Ensemble. Other limitations include retinal layer segmentation errors for the precise OCT en face maps and error introduced through the registration process, where the registered image may be slightly sheared, rotated, or translated compared to the OCT enface image.

In summary, we report in this study multiple methods of incorporating longitudinal data in the prediction of future regions of atrophy in Stargardt disease and AMD. There is high agreement to the manual gradings when predicting the whole region of atrophy, which is further improved when cycling the initial predictions, and incorporating them into the originally prior input longitudinal data. The prediction performance in OCT images is comparable to that using FAF which opens a new, more efficient, and practical door in the assessment of atrophy progression for clinical trials and retina clinics, beyond widely used FAF. These results are highly encouraging for a high-performance interval growth prediction when more frequent or longer-term longitudinal data are available in our clinics. This is a pressing task for our next step as the ongoing research.

## 5 Acknowledgements

This work was supported by the National Eye Institute of the National Institutes of Health under Award Number R21EY029839.

## 6 Author Contributions

Z. M. developed the algorithms, performed experiments, and wrote the main manuscript text. Z.

W. helped with the data conversion, curation, and provided the software support. E. X. and Sophia Xu assisted with image registration and ground truth. I. M. assisted with image registration. S. S. provided clinical background and wrote part of the manuscript. Z. H. cultivated the idea of the development and wrote part of the manuscript.

## 8 Additional Information Use of human participants

All data (both Stargardt and geographic atrophy data) used in this project were de-identified according to the Health and Insurance Portability and Accountability Act Safe Harbor.

For the Stargardt data, the ethics reviews and institutional review board approvals were obtained from the local ethics committees of all the nine participating institutions, i.e., The Wilmer Eye Institute, Johns Hopkins University, Baltimore, Maryland (JHU); Greater Baltimore Medical Centre, Baltimore, Maryland (GBMC); Scheie Eye Institute, University of Philadelphia, Philadelphia, Pennsylvania (PENN); Retina Foundation of the Southwest, Dallas, Texas (RFSW); Moran Eye Centre, Salt Lake City, Utah (MEC); Cole Eye Institute, Cleveland Clinic, Cleveland, Ohio (CC); Moorfields Eye Hospital, London, UK (MEH, UK); Université de Paris 06, Institut national de la santé et de la recherche médicale, Paris, France (INSERM, France); and Eberhard-Karls University Eye Hospital, Tuebingen, Germany (EKU, Germany). Written informed consent from all subjects and/or their legal guardian(s) for both study participation and publication of identifying images were obtained.

For the geographic atrophy data, ethics review and institutional review board approval from the University of California – Los Angeles were obtained. The research was performed in accordance with relevant guidelines/regulations, and informed consent was obtained from all participants.

## Data availability

The datasets generated and/or analyzed during the current study are not publicly available due to the patients’ privacy and the violation of informed consent but are available from the corresponding author on reasonable request.

## References

1. R. W. Strauss et al., “The natural history of the progression of atrophy secondary to Stargardt disease (ProgStar) studies: design and baseline characteristics: ProgStar report no. 1”, Ophthalmology, vol. 123, no. 4, pp. 817–828, 2016. DOI: 10.1016/j.ophtha.2015.12.009.

2. E. M. Schönbach et al., “Macular sensitivity measured with microperimetry in Stargardt disease in the progression of atrophy secondary to Stargardt disease (ProgStar) study: report no.7”, JAMA Ophthalmology, vol. 135, no. 7, p. 696, 2017. DOI: 10.1001/jamaophthalmol.2017.1162.

3. R. W. Strauss et al., “Incidence of atrophic lesions in Stargardt disease in the progression of atrophy secondary to Stargardt disease (ProgStar) study: report no. 5”, JAMA Ophthalmology, vol. 135, no. 7, p. 687, 2017. DOI: 10.1001/jamaophthalmol.2017.1121.

4. R. W. Strauss et al., “Progression of Stargardt disease as determined by fundus autofluorescence in the retrospective progression of Stargardt disease study (ProgStar report no. 9)”, JAMA Ophthalmology, vol. 135, no. 11, p. 1232, 2017. DOI: 10.1001/jamaophthalmol.2017.4152.

5. L. Ma, Y. Kaufman, J. Zhang and I. Washington, “C20-D3-vitamin A slows lipofuscin accumulation and electrophysiological retinal degeneration in a mouse model of Stargardt disease”, Journal of Biological Chemistry, vol. 286, no. 10, pp. 7966–7974, 2010. DOI: 10.1074/jbc.m110.178657.

6. J. Kong et al., “Correction of the disease phenotype in the mouse model of Stargardt disease by lentiviral gene therapy”, Gene Therapy, vol. 15, no. 19, pp. 1311–1320, 2008. DOI: 10.1038/gt.2008.78.

7. K. Binley et al., “Transduction of photoreceptors with equine infectious anemia virus lentiviral vectors: safety and biodistribution of StarGen for Stargardt disease”, Investigative Opthalmology & Visual Science, vol. 54, no. 6, p. 4061, 2013. DOI: 10.1167/iovs.13-11871.

8. N. Mukherjee and S. Schuman, “Diagnosis and management of Stargardt disease”, EyeNet Magazine, American Academy of Ophthalmology, Dec. 2014.

9. X. Kong, A. Ho, B. Munoz, S. West, R. W. Strauss, A. Jha, A. Ervin, J. Buzas, M. Singh, Z. Hu, J. Cheetham, M. Ip, H. P. N. Scholl, “Reproducibility of measurements of retinal structural parameters using optical coherence tomography in Stargardt disease”, Translational Vision Science & Technology 8 (3), 46–46, 2019.

10. Bressler NM, Bressler SB, Congdon NG, Ferris FL 3rd, Friedman DS, Klein R, Lindblad AS, Milton RC, Seddon JM; Age-Related Eye Disease Study Research Group, Potential public health impact of Age-Related Eye Disease Study results: AREDS report no. 11. Arch Ophthalmol. 2003 Nov;121(11):1621–4. PMID: 14609922 PMCID: PMC1473209

11. Davis MD, Gangnon RE, Lee L-Yet al (2005) The age-related eye disease study severity scale for age-related macular degeneration: AREDS report no. 17. Arch Ophthalmol 123:1484–1498

12. Ferris FL, Davis MD, Clemons TE et al (2005) A simplified severity scale for age-related macular degeneration: AREDS report no. 18. Arch Ophthalmol 123:1570–1574

13. Klein R, Klein BE, Jensen SC, Meuer SM. The five-year incidence and progression of age-related maculopathy: the Beaver Dam Eye Study. Ophthalmology. 1997;104(1): 7e21.

14. Blair C. J., “Geographic atrophy of the retinal pigment epithelium: a manifestation of senile macular degeneration”, Arch Ophthalmol. 1975; 93:19–25. PMID: 1111482.

15. S. Schmitz-Valckenberg, F. Holz, A. Bird and R. Spaide, “Fundus autofluorescence imaging: review and persepctives”, Retina, vol. 28, no.3, pp. 385–409, 2008. DOI: 10.1097/iae.0b013e318164a907.

16. D. Huang et al. “Optical coherence tomography”, Science, vol. 254, no. 5035, pp. 1178–1181, 1991. DOI: 10.1126/science.1957169.

17. J. G. Fujimoto et al., “New technology for high-speed and high-resolution optical coherence tomography”, Ann. NY Acad. Sci., vol. 838, no. 1, pp. 96–107, 1998. DOI: 10.1111/j.1749-6632.1998.tb08190.x.

18. Mishra, Z.; Wang, Z.; Sadda, S.R.; Hu, Z. Using Ensemble OCT-Derived Features beyond Intensity Features for Enhanced Stargardt Atrophy Prediction with Deep Learning. Appl. Sci. 2023, 13, 8555. 10.3390/app13148555

19. Z. Mishra, Z. Wang, S. R. Sadda, Z. Hu. “Automatic Segmentation in Multiple OCT Layers For Stargardt Disease Characterization Via Deep Learning”, Translational Vision Science & Technology, vol. 10, no. 4, p. 24, 2021. DOI: 10.1167/tvst.10.4.24.

20. J. Kugelman, D. Alonso-Caneiro, Y. Chen et al. “Retinal boundary segmentation in Stargardt disease optical coherence tomography images using automated deep learning”, Transl Vis Sci Technol, vol. 9, no. 11, p. 12, 2020.

21. J. Charng, D. Xiao, M. Mehdizadeh et al. “Deep learning segmentation of hyperautofluorescent fleck lesions in Stargardt disease”, Sci Rep, vol. 10, p. 16491, 2020.

22. Schmitz-Valckenberg S., Brinkmann C. K., Alten F., Herrmann P., Stratmann N. K., Göbel A. P., Fleckenstein M., Diller M., Jaffe G. J., and Holz F. G., “Semiautomated image processing method for identification and quantification of geographic atrophy in age-related macular degeneration”. Invest Ophthalmol Vis Sci., 2011; 52(10):7640–6.

23. Q. Chen, S. L. de, T. Leng, L. Zheng, L. Kutzscher, and D. L. Rubin, “Semi-automatic geographic atrophy segmentation for SD-OCT images,” Biomed Opt Express 4(12), pp. 2729–50, 2013.

24. S. Wang, Z. Wang, S. Vejalla, A. Ganegoda, M. Nittala, S. Sadda, Z. Hu, Reverse engineering for reconstructing baseline features of dry age-related macular degeneration in optical coherence tomography. Sci Rep 12, 22620 (2022). 10.1038/s41598-022-27140-8. PMID: 36587062. PMCID: PMC9805430

25. Z. Wang, S. R. Sadda, A. Lee, and Z. Hu, Automated segmentation and feature discovery of agerelated macular degeneration and Stargardt disease via self-attended neural networks. Sci Rep 12, 14565 (2022). 10.1038/s41598-022-18785-6. PMID: 36028647. PMCID: PMC9418226

26. Z. tHu, X. Wu, A. Hariri, S. Sadda, “Multiple Layer Segmentation and Analysis in Three-Dimensional Spectral-Domain Optical Coherence Tomography Volume Scans”, J. Biomed. Opt. 0001;18(7):076006–076006. doi:10.1117/1.JBO.18.7.076006. PMID: 23843084.

27. Z. Hu, G. G. Medioni, M. Hernandez, A. Hariri, X. Wu, S. R. Sadda, “Segmentation of the Geographic Atrophy in Spectral-Domain Optical Coherence Tomography and Fundus Autofluorescene Images”, Invest. Ophthalmol. Vis. Sci., December 30, 2013, vol. 54, no. 13, 8375–8383. PMID: 24265015.

28. Z. Wang, S. R. Sadda, and Z. Hu, “Deep learning for automated screening and semantic segmentation of age-related and juvenile atrophic macular degeneration,” in Medical Imaging 2019: Computer-Aided Diagnosis, vol. 10950, International Society for Optics and Photonics, Mar. 2019, 109501Q. DOI: 10.1117/12.2511538.

29. Z. Hu, Z. Wang, S. Sadda, “Automated segmentation of geographic atrophy using deep convolutional neural networks”, Proceedings Volume 10575, SPIE Medical Imaging 2018: Computer-Aided Diagnosis; 1057511 (2018); doi: 10.1117/12.2287001

30. S Saha, Z Wang, S Sadda, Y Kanagasingam, Z. Hu, “Visualizing and understanding inherent features in SD-OCT for the progression of age-related macular degeneration using deconvolutional neural networks”, Applied AI Letters, vol. 1, no. 1, 2020. 10.1002/ail2.16

31. U. Schmidt-Erfurth, H. Bogunovic, C. Grechenig, P. Bui, M. Fabianska, S. Waldstein, and G. S. Reiter, “Role of Deep Learning-Quantified Hyperreflective Foci for the Prediction of Geographic Atrophy Progression,” eng, American Journal of Ophthalmology, vol. 216, pp. 257–270, Aug. 2020, ISSN: 1879-1891.DOI: 10.1016/j.ajo.2020.03.042.

32. D. Ramsey, J. Sunness, P. Malviya, C. Applegate, G. Hager, and J. Handa, “Automated image alignment and segmentation to follow progression of geographic atrophy in age-related macular degeneration,” Retina 34, pp. 1296–307, 2014.

33. Bart Liefers, Johanna M. Colijn, Cristina González-Gonzalo, Timo Verzijden, Jie Jin Wang, Nichole Joachim, Paul Mitchell, Carel B. Hoyng, Bram van Ginneken, Caroline C.W. Klaver, Clara I. Sánchez, A Deep Learning Model for Segmentation of Geographic Atrophy to Study Its Long-Term Natural History, Ophthalmology, Volume 127, Issue 8, 2020, Pages 1086–1096, ISSN 0161-6420, 10.1016/j.ophtha.2020.02.009.

34. Chu Z, Wang L, Zhou X, Shi Y, Cheng Y, Laiginhas R, Zhou H, Shen M, Zhang Q, de Sisternes L, Lee AY, Gregori G, Rosenfeld PJ, Wang RK. Automatic geographic atrophy segmentation using optical attenuation in OCT scans with deep learning. Biomed Opt Express. 2022 Feb 7;13(3):1328–1343. doi: 10.1364/BOE.449314. PMID: 35414972; PMCID: PMC8973176.

35. Pramil V, de Sisternes L, Omlor L, Lewis W, Sheikh H, Chu Z, Manivannan N, Durbin M, Wang RK, Rosenfeld PJ, Shen M, Guymer R, Liang MC, Gregori G, Waheed NK. A Deep Learning Model for Automated Segmentation of Geographic Atrophy Imaged Using Swept-Source OCT. Ophthalmol Retina. 2023 Feb;7(2):127–141. doi: 10.1016/j.oret.2022.08.007. Epub 2022 Aug 12. PMID: 35970318.

36. Kalra, G.; Cetin, H.; Whitney, J.; Yordi, S.; Cakir, Y.; McConville, C.; Whitmore, V.; Bonnay, M.; Lunasco, L.; Sassine, A.; Borisiak, K.; Cohen, D.; Reese, J.; Srivastava, S.K.; Ehlers, J.P. Machine Learning-Based Automated Detection and Quantification of Geographic Atrophy and Hypertransmission Defects Using Spectral Domain Optical Coherence Tomography. J. Pers. Med. 2023, 13, 37. 10.3390/jpm13010037

37. Ji Z, Chen Q, Niu S, Leng T, Rubin D. L., “Beyond Retinal Layers: A Deep Voting Model for Automated Geographic Atrophy Segmentation in SD-OCT Images”, Transl Vis Sci Technol. 2018 Jan 2; 7(1):1. doi: 10.1167/tvst.7.1.1. eCollection 2018 Jan. PMCID: PMC5749649. PMID: 2930238

38. Manaswi, N.K. (2018). RNN and LSTM. In: Deep Learning with Applications Using Python. Apress, Berkeley, CA. 10.1007/978-1-4842-3516-4_9

39. Calin, Ovidiu (14 February 2020). Deep Learning Architectures. Cham, Switzerland: Springer Nature. p. 555. ISBN 978-3-030-36720-6.

40. A. Graves, J. Schmidhuber. Offline Handwriting Recognition with Multidimensional Recurrent Neural Networks. NIPS’22, p 545–552, Vancouver, MIT Press, 2009.

41. Kugelman, J. Automatic segmentation of OCT retinal boundaries using recurrent neural networks and graph search. Biomedical optics express, 9(11), 5759–5777.

42. Hochreiter, S. & Schmidhuber, Juergen, 1997. Long short-term memory. Neural computation, 9(8), pp.1735–1780

43. Monner D, Reggia JA. A generalized LSTM-like training algorithm for second-order recurrent neural networks. Neural Netw. 2012 Jan;25(1):70–83. doi: 10.1016/j.neunet.2011.07.003. Epub 2011 Jul 18. PMID: 21803542; PMCID: PMC3217173.

44. Avyuk Dixit, Jithin Yohannan, Michael V. Boland, Assessing Glaucoma Progression Using Machine Learning Trained on Longitudinal Visual Field and Clinical Data, Ophthalmology, Volume 128, Issue 7, 2021, Pages 1016–1026, ISSN 0161-6420.

45. Lee, J. et al. (2022). Predicting Age-related Macular Degeneration Progression with Longitudinal Fundus Images Using Deep Learning. In: Lian, C., Cao, X., Rekik, I., Xu, X., Cui, Z. (eds) Machine Learning in Medical Imaging. MLMI 2022. Lecture Notes in Computer Science, vol 13583. Springer, Cham. 10.1007/978-3-031-21014-3_2

46. Santeramo, R., Withey, S., & Montana, G. (2018). Longitudinal detection of radiological abnormalities with time-modulated LSTM. In Deep Learning in Medical Image Analysis and Multimodal Learning for Clinical Decision Support: 4th International Workshop, DLMIA 2018, and 8th International Workshop, ML-CDS 2018, Held in Conjunction with MICCAI 2018, Granada, Spain, September 20, 2018, Proceedings 4 (pp. 326–333). Springer International Publishing.

47. Hong, X., Lin, R., Yang, C., Zeng, N., Cai, C., Gou, J., & Yang, J. (2019). Predicting Alzheimer’s disease using LSTM. Ieee Access, 7, 80893–80901.

48. M. Hernandez, G. G. Medioni, Z. Hu, S. R. Sadda, “Multimodal registration of multiple retinal images based on line structures”, Applications of Computer Vision (WACV), 2015 IEEE Winter Conference on 5-9 Jan. 2015, pp. 907–914.

